# Protection against infection with the Omicron BA.5 subvariant among people with previous SARS-CoV-2 infection - surveillance results from southern Sweden, June to August 2022

**DOI:** 10.1101/2022.11.08.22282069

**Authors:** Fredrik Kahn, Carl Bonander, Mahnaz Moghaddassi, Louise Bennet, Ulf Malmqvist, Malin Inghammar, Jonas Björk

## Abstract

We evaluated the protection afforded by SARS-CoV-2 natural infection against reinfection among vaccinated during a calendar period from June to August 2022 when Omicron BA.5 was the dominating subvariant in Scania county, Sweden. We formed a study cohort (n = 71 592) mainly consisting of health care workers by restricting to people 18-64 years old who received their first vaccine dose relatively early (24 April 2021 or sooner). We used continuous density case-control 1:10 sampling matched for sex and age within the study cohort, and thereby obtained 1 114 cases during Omicron BA.5 dominance and 11 140 controls who were analysed with conditional logistic regression. Limited protection against reinfection was suggested from prior infection of virus variants before Omicron (11%, 95% confidence interval [CI] −10 to 28%]. By contrast, prior Omicron infection offered clear protection (65%, 95% CI 56-73%). For the Omicron BA.2 subvariant, stronger protection was suggested during early (85%, 95% CI 75-91%) than later BA.5 dominance (66%, 95% CI 48-78%). Lower protection was observed from the previous BA.1 subvariant (30%; 95% CI −4 to 53%). These findings suggest that natural infection from the Omicron subvariants contributes to short-term population protection against reinfection with the subvariant BA.5 among vaccinated, but wanes considerably 5-6 months after infection.

---

The severe acute respiratory syndrome coronavirus 2 (SARS-CoV-2) variant of concern (VOC) Omicron (Phylogenetic Assignment of Named Global Outbreak (Pango) lineage designation B.1.1.529) currently dominates by the subvariant BA.5 in many countries. It is important to study the protection from prior infection with the subvariants BA.1 and BA.2 against BA.5 infection as some of the adapted vaccines in clinical trials are based on BA.1 (1). A recent study from a highly vaccinated study population in Portugal reported 75% protection from prior BA.1/BA.2 infection against BA.5 infection (2). The study was large and the statistical precision in the estimated protection was therefore high but the follow-up period was short. Similar protection from prior BA.1/BA.2 infection on BA.4/BA.5 infection, but with much wider confidence intervals, has also been observed in a study from Qatar (3). Protection exceeding 90% from BA.1/BA.2 has been reported in a Danish study, using test-negative controls (4), but their follow-up period with BA.5 dominance was relatively short, and waning immunity could not be assessed. Rapidly waning protection induced by prior BA.1/BA.2 infection has been reported from Portugal when contrasting reinfection risks three and five months later (5). There is a need for additional studies with longer follow-up to investigate the length of protection. The present study aimed to evaluate protection afforded by SARS-CoV-2 natural infection against reinfection with the Omicron BA.5 subvariant among vaccinated. The study was conducted in Scania county, southern Sweden, a region that had a rapid transition from previous Omicron variants to BA.5.

## Methods

### Study design and data extraction

The overall study population included all persons residing in Scania county (Skåne), southern Sweden, on 27 December 2020 (baseline) when vaccinations started (n = 1 384 531) (6, 7). Data from national and regional register holders were linked using the personal identification number assigned to all Swedish residents (8). Weekly updates on vaccination date, type of vaccine and dose were obtained from the National Vaccination Register, and data on COVID-19 cases (defined by a positive SARS-CoV-2 test result) from the electronic system SMINet, both kept at the Public Health Agency of Sweden. Regional health registers were used as complementary data sources to provide data on positive tests rapidly, and to assess comorbidities and disease outcomes.

Comorbidities were defined from diagnoses in inpatient or specialised care at any time point during the five years before baseline in the following disease groups (see Supplementary Table S1 for a detailed list): cardiovascular diseases, diabetes or obesity, kidney or liver diseases, respiratory diseases, neurological diseases, cancer or immunosuppressed states, and other conditions and diseases (Down syndrome, HIV, sickle cell anaemia, drug addiction, thalassaemia or mental health disorder). The number of comorbidities in these groupings was counted.

From the overall study population, we formed a cohort (n = 71 592) restricted to working-age people (18-64 years old) who received their first vaccine dose relatively early (24 April 2021 or sooner). With these restrictions, we expect the study cohort to mainly consist of health care workers as they were prioritized in the vaccination program (see also *Sensitivity analysis* below). Health care workers were recommended to undergo testing throughout the study period in case of SARS-CoV-2 symptoms. The cohort was followed longitudinally for positive SARS-CoV-2 tests until 9 August 2022 (week 32). Individuals who died or moved away from the region were censored on the date of death or relocation. The follow-up period was grouped in accordance with routine sequencing of samples of infected cases in Scania county on dominating VOC (cf. Supplementary Figure S1): i) before Omicron (until 2021 week 47), ii) transition to Omicron (2021 week 48-51), iii) Omicron BA.1 dominance (60%; 2021 week 52 until 2022 week 1), iv) transition to Omicron BA.2 (2022 week 2-3), v) Omicron BA.2 dominance (89%; 2022 week 4-20), vi) transition to Omicron BA.5 (2022 week 21-24), vii) Omicron BA.5 dominance (86%; 2022 week 25-32). In this study, we focused specifically on the follow-up period starting in 2022 week 25 (20 June) when Omicron BA.5 started to be the dominating VOC (>70%) and assessed the protective effect of the latest prior infection overall and during early (week 25-28) and late (29-32) follow up.

### Statistical analysis

We used continuous density case-control sampling (9) nested within the study cohort. For each infected case during follow-up, ten controls without a positive test the same week as the case or 90 days prior were randomly selected from the study cohort, matched with respect to sex and age (five-year groups). Using conditional logistic regression, we estimated protection against infection with the Omicron BA.5 subvariant associated with the timing of the latest previous SARS-CoV-2 infection grouped according to the description above and adjusted for the number of vaccine doses. Only vaccine doses obtained seven or more days before the case date were counted in the analyses. BNT16b2 mRNA (Comirnaty, Pfizer-BioNTech) was the most frequently used vaccine type with 65% of the 208 651 administrated doses in the study cohort. Protection was reported as 1 – OR (odds ratio).

### Sensitivity analysis

As early vaccination among working-age people in Sweden was not only offered to health-care personnel but also to specific risk groups we also conducted two sensitivity analyses to assess the robustness of the findings: i) excluding individuals with organ or hematopoietic stem cell transplantation, undergoing dialysis or with a third vaccine dose obtained early (28 September 2021 or sooner; before third dose was offered to health-care personnel), ii) updated case-control sampling after restricting the study cohort further to individuals without any comorbidity (n = 54 187).

## Results

A total of 1 114 COVID-cases occurred during the period with Omicron BA.5 dominance, who were used to sample 11 140 matched controls (Table 1). Case and controls were similar with respect to civil status, proportion born abroad and number of comorbidities. No protection from booster vaccine doses was observed; on the contrary the proportion who had received at most two vaccine doses was higher among controls than among cases (12.7 vs. 8.5%). The risk of hospitalisation was low among the confirmed cases during the period (1.6%; 18 out of 1 114 cases), albeit higher than during the period with BA.1 dominance (0.5%; 12 out of 2 501 cases).

**Table 1.** Characteristics of the COVID-19 cases (N = 1 114) and sex and age matched controls (N = 11 140) during the follow up period in June – August 2022 when Omicron BA.5 was the dominating SARS-CoV-2 subvariant in Scania county, Sweden

|  | Cases, n(%) |  | Controls, n(%) |  |
| --- | --- | --- | --- | --- |
| Total | 1 114 (100) |  | 11 140 (100) |  |
| Age |  |  |  |  |
| 18 – 34 | 290 (26) |  | 2 825 (25) |  |
| 35 – 44 | 203 (18) |  | 2 182 (20) |  |
| 45 – 54 | 287 (26) |  | 2 745 (25) |  |
| ≥ 55 years | 334 (30) |  | 3 388 (30) |  |
| Sex |  |  |  |  |
| Females | 888 (80) |  | 8 880 (80) |  |
| Males | 226 (20) |  | 2 260 (20) |  |
| Born abroad | 207 (19) |  | 2 257 (20) |  |
| Civil status |  |  |  |  |
| Married | 464 (42) |  | 4 600 (41) |  |
| Divorced | 139 (12) |  | 1 530 (14) |  |
| Single | 507 (46) |  | 4 896 (44) |  |
| Widow/widower | 4 (0.4) |  | 114 (1.0) |  |
| Comorbidities |  |  |  |  |
| 0 | 847 (76.0) |  | 8 576 (77.0) |  |
| 1 | 192 (17.2) |  | 1 868 (16.8) |  |
| ≥ 2 | 75 (6.7) |  | 692 (6.2) |  |
| Vaccine doses |  |  |  |  |
| 1 | 6 (0.5) |  | 73 (0.7) |  |
| 2 | 89 (8.0) |  | 1 312 (12) |  |
| 3 | 990 (89) |  | 9 347 (84) |  |
| 4-5 | 29 (2.6) |  | 408 (3.7) |  |
| Prior SARS-CoV-2 infection | <u>Months<sup>a</sup></u> | <u>n(%)</u> | <u>Months<sup>a</sup></u> | <u>n(%)</u> |
| No | - | 823 (74) | - | 6 738 (60) |
| Before Omicron | 19 | 170 (15) | 19 | 1 569 (14) |
| Transition to Omicron | 7.0 | 7 (0.6) | 7.0 | 144 (1.3) |
| Omicron BA.1 | 6.4 | 33 (3.0) | 6.4 | 393 (3.5) |
| Transition to BA.2 | 6.1 | 37 (3.3) | 5.9 | 757 (6.8) |
| Omicron BA.2 | 5.8 | 44 (3.9) | 5.2 | 1 539 (14) |

<sup>a</sup>Median time since prior infection in months

Limited protection against reinfection was suggested from prior infection of virus variants before Omicron (11%, 95% confidence interval [CI] −8 to 28%). By contrast, prior infection with Omicron offered clear protection against BA.5 (66%; 95% CI 56-73%). This protection was similarly high among those with two prior infections (one before and one during Omicron dominance; 74%; 95% CI 52-86%). In sensitivity analyses, excluding risk group individuals who were vaccinated early (30 cases and 244 controls), or restricting the case-control sampling to individuals without comorbidities (847 cases and 8 470 controls), did not alter the estimated protection associated with prior Omicron infection (67%, 95% CI 58-74%, and 69%, 95% CI 59-76%, respectively).

Among the Omicron subvariants, stronger protection was observed for the more recent variant BA.2 (77%; 95% CI 68-83%; median 5.2 months after infection among controls) than for BA.1 (30%; 95% CI −4 to 53%; median 6.4 months; Figure 1). The protection associated with prior BA.2 infection was strong during the early period of BA.5 dominance (85%, 95% CI 75-91%; 4.9 months in median since prior infection among controls), whereas the protection during the later BA.5 period was moderate (66%, 95% CI 48-78%; 5.8 months in median) and similar to the average protection afforded from prior infection during the transition period from BA.1 to BA.2 (60%, 95% CI 42-72%; 5.9 months in median since prior infection).

**Figure 1.**
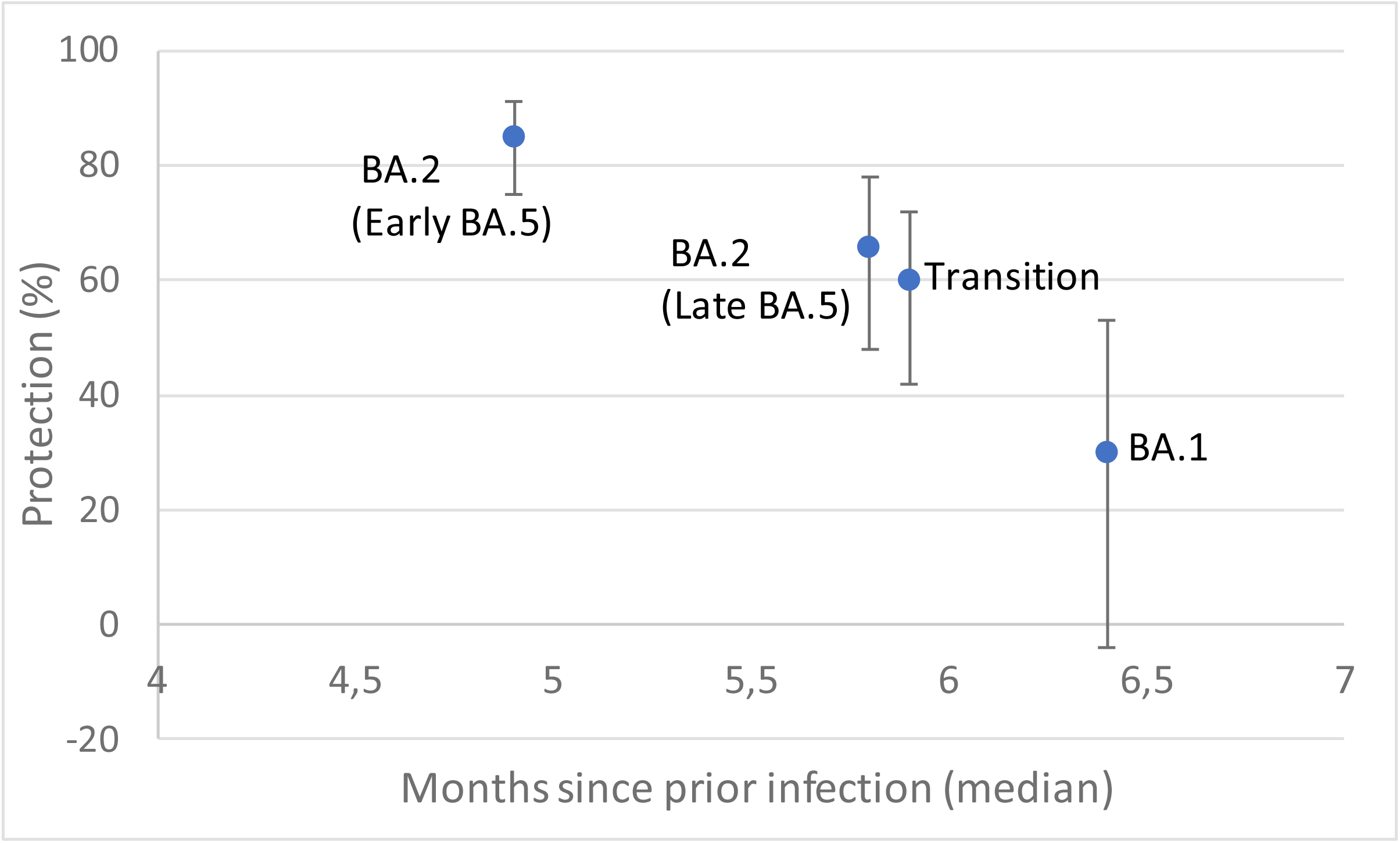
Protection (%) against SARS-CoV-2 reinfection with the Omicron BA.5 subvariant in relation to average time since prior infection and dominating variant at time of prior infection: BA.2, transition from BA.1 or BA.1. Results for prior BA.2 infection are further stratified on early and late follow up period with BA.5 dominance.

## Ethical statement

Ethical approval was obtained from the Swedish Ethical Review Authority (2021-00059).

## Discussion

A salient finding of the present study among working-age vaccinated was the marked short-term protection against Omicron BA.5 infection associated with prior infection with previous Omicron subvariants. A previous population study from Sweden with follow-up that ended before Omicron found that the relative risk of SARS-CoV-2 reinfection in individuals who survived a previous infection remained low for up to 20 months (10). The emergence of Omicron has markedly shortened the duration of the protection (5). Our study adds important new evidence in that respect by i) having a longer follow-up with Omicron dominance compared with previous studies (2-4) and ii) being able to stratify the protection further by Omicron subvariants. Although the protection against reinfection in the present study was mainly observed among persons previously infected with the BA.2 subvariant, differences in time since prior infection with BA.1 and BA.2 should be considered. We found similar protection associated with prior BA.2 infection during the later follow-up period as the average protection for the complete follow up associated with prior infection during the transition period from BA.1 to BA.2. The average time since prior infection was also similar in this comparison, which suggests that the protection afforded by BA.2 remains as long as for BA.1. The protection against reinfection remained stable 5 months after infection and then waned considerably irrespectively of subvariant, which is in line with another recent study on durability of immune protection (5). Thus, a booster dose seems unnecessary until 5 months after a confirmed infection.

We have previously reported for this study population how the protection from two vaccine doses against infection disappeared after the emergence of Omicron (7). A secondary finding of the present study was that the first booster dose also did not generate detectable protection against infection in the population. This is consistent with a small immunological study with repeated collection of sera and nasal swabs in 27 individuals that showed that infection, but not vaccination, triggered detectable local response against SARS-CoV-2 in the nasal mucosa (11). Furthermore, another immunological study has found that BA.4 and BA.5 were less susceptible than BA.1 and BA.2 to vaccine elicited antibody neutralization (12).

We found higher risk of hospitalisation among confirmed cases during BA.5 than previously during BA.1 dominance. A similar increase in risk of hospitalisation was noted in a Danish study when contrasting BA.2 and BA.5 infections (4), but increasing selection in the confirmed cases during follow-up is an alternative explanation for these findings. It was not possible to assess changes vaccine protection against hospitalisation in our low-risk cohort. A recent study from England found no evidence of decreasing vaccine effectiveness 2-14 weeks after a booster dose against hospitalisation for BA.4 or BA.5 as compared to BA.2 (13).

We assessed the protection from two prior infections in a secondary analysis, because a previous study has suggested that the immune boosting by Omicron is lost with prior Wuhan-Hu-1 imprinting (14). However, that hypothesis could not be confirmed in our study as we saw similar protection in individuals with prior Omicron infection irrespectively whether they had another confirmed infection before Omicron.

The key strength of our study was the detailed individual-level data on vaccinations and infections during the entire study period. A major limitation was that we only had data on dominating virus variants at the population-level at different periods and not for individual cases. Our study may therefore have underestimated the difference in natural protection from different virus variants. Undetected current or prior infections at the time of control sampling due to limited testing may also have biased the estimates towards no protection. The short periods of BA.1 and BA.2 dominance hampered the possibility to separate general waning of immunity from differential protection across these subvariants. It should also be noted that even though our follow-up period was longer than previous studies no person in the cohort had time since prior Omicron infection exceeding seven months. Continued monitoring of natural protection associated with the Omicron subvariants is therefore warranted.

## Conclusion

Natural infection from the Omicron subvariants contributes to short-term population protection against reinfection with the subvariant BA.5, but wanes considerably 5-6 months after infection. No protection from booster vaccine doses against infection was observed in this cohort of working-age vaccinated people.

## Data Availability

All data produced in the present study are available upon reasonable request to the authors, but would generally require a new ethical approval.

## Acknowledgments

Magnus Rasmussen for fruitful discussions when planning the study. Cecilia Åkesson-Kotsaris, Paul Söderholm and Helena Hallefjord, Clinical Studies Sweden, for excellence in bringing the surveillance infrastructure in place. Claus Bohn Christiansen, Scania County Council, Clinical microbiology, for providing data from routine sequencing of infected cases.

## Authors’ contributions

JB and FK conceived the study, with important contributions from CB, MM, LB, UM and MI. UM, JB and MM acquired data, CB, JB and MM conducted the statistical analyses. JB drafted the manuscript with assistance from FK and MI. All authors contributed with interpretation of results, critically revised the manuscript and approved the final version for submission.

