## Supplementary Material for "Protection against infection with the Omicron BA.5 subvariant among people with previous SARS-CoV-2 infection - surveillance results from southern Sweden, June to August 2022"

**Content**

Supplementary Table S1
Supplementary Figure S1

**Supplementary Table S1.** Classification of comorbidities.

| **Disease group** | **ICD-10 codes (incl KVÅ-codes[a])** |
|---|---|
| Cardiovascular diseases | I10-I15, I20-I25, I42-I43<br>I50, I60-I69<br>J81 |
| Diabetes or obesity | E10, E11, E66 |
| Kidney or liver diseases | K70.X, K74.3-K74.6, K75.4, K76.0<br>N18.5, N18.9<br>DR016, DR024 |
| Respiratory diseases | A15-A19<br>E84<br>I26, I27<br>J42, J43, J44, J45, J47, J84<br>J96, J98.2, J98.3 |
| Neurological diseases (including dementia) | G00-G99<br>F00-F03 |
| Cancer or immunosuppressed state (including organ transplantation) | C00-C99<br>KAS, FQA, FQB, JJC, GDG, JLE<br>DR046, DR047, DR048<br>D80.0-D80.1<br>D80.5, D81, D82, D83 |
| Other conditions and diseases<br>• HIV<br>• Thalassemia<br>• Sickle cell<br>• Mood disorders<br>• Schizophrenia spectrum disorders<br>• Substance use disorders<br>• Downs syndrome | B20-B24<br>D56, D57<br>F10-F19, F30-F39, F20-F29<br>Q90 |

[a] Swedish classification of certain interventions during health care visits

**Supplementary Figure S1.** Routine sequencing of samples of infected cases in Scania county, Sweden during the follow-up period with Omicron dominance 2021, week 52 – 2022 week 32.

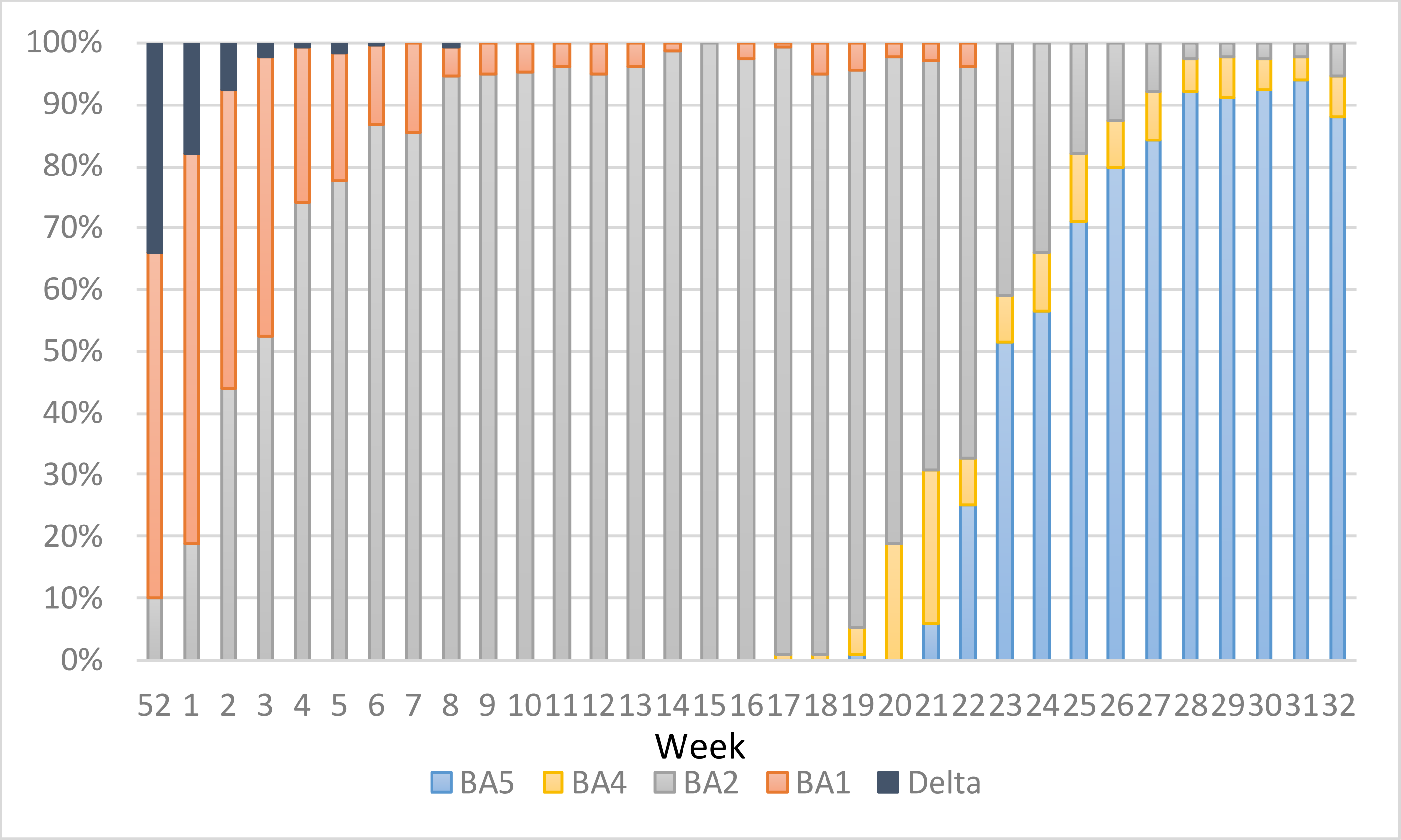
